# Durable Tumor Volume Plateau Characterizes Long-Term Responders to Ivosidenib in IDH-Mutant Glioma

**DOI:** 10.64898/2026.11.26.26368351

**Authors:** Sushant Puri, Lindsey Greenlund, Dylan Ross, Peng Huang, Sriya Pokala, Ruth Agyekum, David O Kamson, Calixto Hope-Lucas, Kristin Redmond, Haris Sair, Lawrence R Kleinberg

## Abstract

**Purpose/Objectives:** IDH inhibitor therapy is now FDA-approved for low-grade IDH-mutated (mIDH) gliomas. Early studies with short follow-up periods have reported variable tumor volume reduction. We describe radiographic responses in long-term responders.

**Materials/Methods:** MRIs in patients with progressive mIDH gliomas and subsequently treated with targeted therapy for >36 months from 2019-2024 were retrospectively reviewed. Contouring of T2/FLAIR sequences were performed to calculate tumor volumes.

**Results:** From a total of 49 consecutive patients started on targeted therapy, 13 were included (11 astrocytoma, 2 oligodendroglioma). A total of 157 MRIs obtained prior to and during ivosidenib were analyzed. Median duration of treatment was 59.5 months (range 37.4-79.3) with 1/13 offered chemoradiation due to radiographic progression. Pretreatment tumor volumes increased at the average rate of 12.6 cc/year (p = 0.0007) and tumor volumes declined during the first year of therapy at an average rate of −9.73 cc/year (p < 0.001). After one year of therapy, tumor volume changes were no longer significantly different from zero (p = 0.36).

**Conclusions:** Our real-world data demonstrates a reduction in tumor volume during the first year of treatment of progressive mIDH glioma with ivosidenib followed by prolonged stabilization. The principal benefit of mutant IDH inhibition may not be prolonged tumor volume reduction but durable stable disease. Our findings can inform clinical practice using targeted therapy over an extended period which is currently unreported.

**Key points:**

1. Early tumor volume reduction is followed by durable radiographic stability.
2. One in four patients remained on targeted therapy beyond 3 years.
3. Long-term disease control may defer chemoradiation in selected patients.

## Introduction

Mutations in isocitrate dehydrogenase (IDH) define a distinct molecular class of diffuse gliomas, differentiating them from IDH wild-type glioblastoma [1-3]. IDH mutated (mIDH) gliomas are divided into astrocytoma or oligodendroglioma. These are relatively rare with an annual incidence of 0.44/100,000 individuals with approximately 3000 cases newly identified in the United States annually [4]. Treatment of mIDH gliomas includes maximal safe surgical resection followed by observation or adjuvant radiotherapy and chemotherapy depending on patient and disease risk factors [5, 6]. Lower-grade (WHO grade 2 and 3) mIDH gliomas often have prolonged clinical courses, affecting patients during their most productive years hence are associated with a significantly higher Disability-Adjusted Life Years (DALY) and treatments affecting Health Related Quality of Life (HRQOL)[7].

Mutation in the IDH enzyme leads to an accumulation of oncometabolite, 2-hydroxyglutarate (2-HG) that results in widespread epigenetic and metabolic changes, as well as local immune suppression [8]. 2HG is also epileptogenic and is responsible for seizures that affect the quality of life and outcomes [9-11]. These biological findings have led to the development of mutant IDH inhibitors (mIDHi) as a safe and effective therapeutic strategy. [12-14]. Ivosidenib, a first-in-class inhibitor of IDH1 with blood-brain-barrier permeability has demonstrated a satisfactory safety profile and delaying Progression Free Survival (PFS) [12, 15-17]. More recently, INDIGO (INvestigating vorasiDenIb in GliOma: NCT04164901) evaluated vorasidenib, a dual IDH1/IDH2 inhibitor, in patient with recurrent WHO Grade 2 mIDH gliomas without prior radiation or chemotherapy. Vorasidenib significantly improved PFS and Time to Next Intervention (TTNI) compared with placebo [13, 18]. It also led to an improvement in seizure control without detrimental effects on HRQOL or cognition [18]. Based on these results vorasidenib, is now FDA approved for WHO Grade 2 non-enhancing, recurrent, IDH mutated gliomas [19].

As mIDH inhibitors are incorporated into routine clinical practice, patterns of treatment response over extended periods is of increasing importance. Conventional radiographic response assessment in non-enhancing gliomas has historically relied on two-dimensional measurements [20]; however, these approaches are limited by irregular tumor morphology, anisotropic growth patterns, residual treatment-related changes, and variability in image acquisition [21-23]. Three-dimensional volumetric assessment provides a more sensitive and reproducible approach for evaluating tumor growth dynamics [24-27]. Three-dimensional measurements have demonstrated improved inter-reader agreement with lower measurement variability compared to two-dimensional methods in mIDH gliomas [24-30].

Prior studies including ours have demonstrated early reductions in tumor volume in up to 75% of patients with WHO Grade 2/3 mIDH gliomas treated with ivosidenib or vorasidenib; however, the long-term clinical and imaging responses beyond early follow-up from clinical trials remains largely unknown [18, 31]. We asked if early responders to IDHm inhibition experienced continued tumor volume reduction, durable stabilization, or alternative patterns of response.

Addressing this knowledge gap is crucial for interpreting clinical benefit and imaging endpoints thus informing treatment decisions in this population. In this study, we evaluated radiographic features from a cohort of patients with recurrent mIDH gliomas treated with targeted therapy, focusing on characterizing long-term, longitudinal changes in tumor volume as imaging-based biomarkers of treatment response.

## Material and Methods

### Patient Selection

This is a single-institution, retrospective study approved by the Johns Hopkins IRB Committee with a waiver of informed consent. This study was performed in compliance with Health Insurance Portability and Accountability Act (HIPAA) regulations as well as the Declaration of Helsinki.

### Inclusion criteria and Exclusion criteria

Patients for this study were screened from the treatment team’s clinical and were also included in our prior study on early imaging kinetics. [31]. Demographic data, pathology and molecular information, treatment history, and imaging data were collected from patient charts. To address our research question of long-term imaging outcomes, we screened consecutive patients with progressive WHO Grade 2 or 3 non-contrast enhancing astrocytoma or oligodendroglioma started on targeted therapy with either ivosidenib or vorasidenib. Vorasidenib was obtained prior to the FDA approval and outside of clinical trials as a single patient IIT. All patients had pathology proven IDH mutated glioma with mutation status confirmed by either immunohistochemistry or next generation sequencing. Tumor subtypes were classified using the 2021 WHO Classification of Tumors of the Central Nervous System. Additional inclusion criteria included (i) pretreatment scans for a duration of at least 12 months and/or 2 or more time points and (ii) more than 36 months follow up. Clinical responses to therapy were initially judged by treating oncologists and radiologists in consultations with multi-disciplinary tumor board discussions if indicated. The short-term imaging dynamics of the patient population have been previously reported [18, 31].

### MRI Acquisition

MRI scans were obtained either 1.5 or 3.0 Tesla scanners using a pre-defined institutional protocol. Conventional MRI sequences included pre- and post-contrast T1-weighted images and 2D or 3D T2-weighted FLAIR images. MRI slice thickness was no more than 5 mm with an interslice gap of no more than 1.5 mm. These were performed as part of routine clinical management. All MRI scans were reviewed independently by radiologist (LG), neurologist (SP), and radiation oncologist (DR).

### Tumor Segmentation, Contouring, and Volumetrics

For all patients included in this study, manual 3D segmentation was performed on Raystation version 2023b from Research Laboratories (DR) and independently reviewed for each patient (LG) in December 2025. The segmented contours excluded nontumor regions which were either surgical cavity, postoperative changes, or hemorrhagic changes. After 3D segmentation was completed, volume in cubic centimeters was automatically calculated with integrated, automated functions within Raystation. Tumor growth rate (TGR) was calculated as the change in tumor volume per year (cc/year).

TGRs were calculated for 12-month time intervals before and after starting mIDHi therapy. Post-treatment TGR were calculated for the entirety of the duration of treatments.

### 3-Dimensional response assessment

Based on previously published criteria for volumetric response assessment in gliomas, we defined the following thresholds (i) Complete response: 100% decrease, (ii) partial response: > 65% decrease, (iii) minor response: between > 65% decrease and <40% decrease, (iv) stable disease: between <40% decrease and <40% increase, and (v) progressive disease: >40% increase **(Supplementary Table 1)** [30].

### Statistical Analysis

Descriptive statistics including proportions, medians, and interquartile ranges (IQR) were used to describe the cohort characteristics. Demographic, tumor and clinical characteristics were analyzed. The primary objective was to characterize longitudinal imaging trajectories rather than identify predictors of response. We used tumor growth rate (TGR) to quantify treatment efficacy in our patient cohort. Accordingly, mixed-effects regression was chosen to estimate average within-patient changes over time while accounting for repeated measurements and unequal follow-up intervals. Time was modeled as a three-segment piecewise linear function to allow for different phases of tumor behavior: (1) the pretreatment period (time < 0 years), (2) the first year after treatment initiation (0–1 year), and (3) the period beyond one year after treatment (>1 year). Three distinct time variables were constructed accordingly: time_pre, representing pretreatment follow-up; time_early, representing the first year on treatment; and time_late, representing follow-up beyond one year. One year was selected a priori based on our previous observation that most radiographic response occurs within the first year mIDHi therapy initiation [31]. Separate mixed-effects models were fitted for tumor volumes. Each model included the three piecewise time variables as fixed effects and a patient-specific random intercept to account for within-subject correlation. Model parameters were estimated using restricted maximum likelihood. Regression coefficients represent the average rate of change within each time interval. Two-sided P values <0.05 were considered statistically significant. Ninety-five percent confidence intervals (95% CIs) were calculated for all fixed-effect estimates. Because the primary objective was to characterize overall longitudinal imaging trajectories, our models were not adjusted for additional covariates. Statistical analyses were performed in R (R Foundation for Statistical Computing, Vienna, Austria).

## Results

### Summary of the study patient population

Between January 2019 and June 2024, 49 patients with pathology confirmed recurrent mIDH gliomas (astrocytoma or oligodendroglioma) were treated with mIDHi (ivosidenib or vorasidenib). All patients had measurable progressive tumor tumor prior to initiating mIDHi therapy; 41 patients were radiation/chemo naïve and eight patients had received prior radiation + temozolomide. 46 were treated off-label with ivosidenib and 3 received vorasidenib **(Fig. 1)**. Of these 49 patients, 13 progressed who were offered alternative therapies and are excluded from this analysis. Notably, all treatment failures occurred in the first two years of treatment: 6/13 (46.1%) within the first year and 7/13 (53.9%) between the first and second years. 3 patients died due to progressive disease after treatment failure. 34 patients remained on treatment at the time of analysis with disease control. All patients were on treatment for at least one year: 11 patients were on treatment between 12 and 24 months, 10 patients between 24 and 36 months, 5 patients between 36 and 48 months, 2 patients between 48 and 60 months, 3 patients between 60 and 72 months, and 3 patients over 72 months.

**Figure 1.**
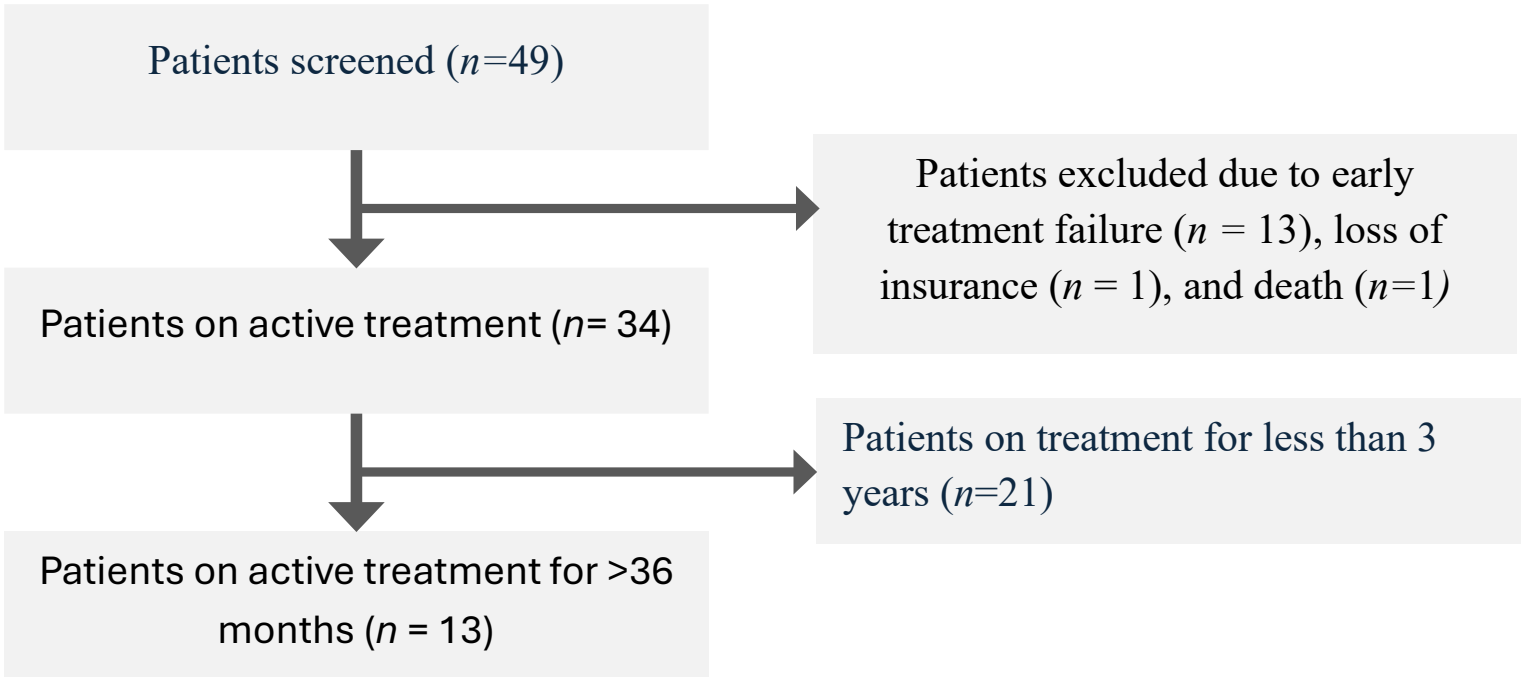
Study Schematic. The 13/49 (26.5%) patients who continued mIDH therapy for over 36 months are the focus of this study. **(Fig. 2)**. In this final cohort, the median age was 38 years (range 26-56 years). The diagnosis was astrocytoma in 11/13 patients (84.6%) (9 WHO grade 2 and 2 WHO grade 3) and 2/13 had oligodendroglioma (both grade 2) (15.3%). 12/13 (92%) patients were male. Median duration of treatment was 59.5 months (range 37.4 to 79.3) **(Table 1)**. No patients on mIDHi therapy for over 36 months paused therapy for toxicity or other reasons.

**Figure 2.**
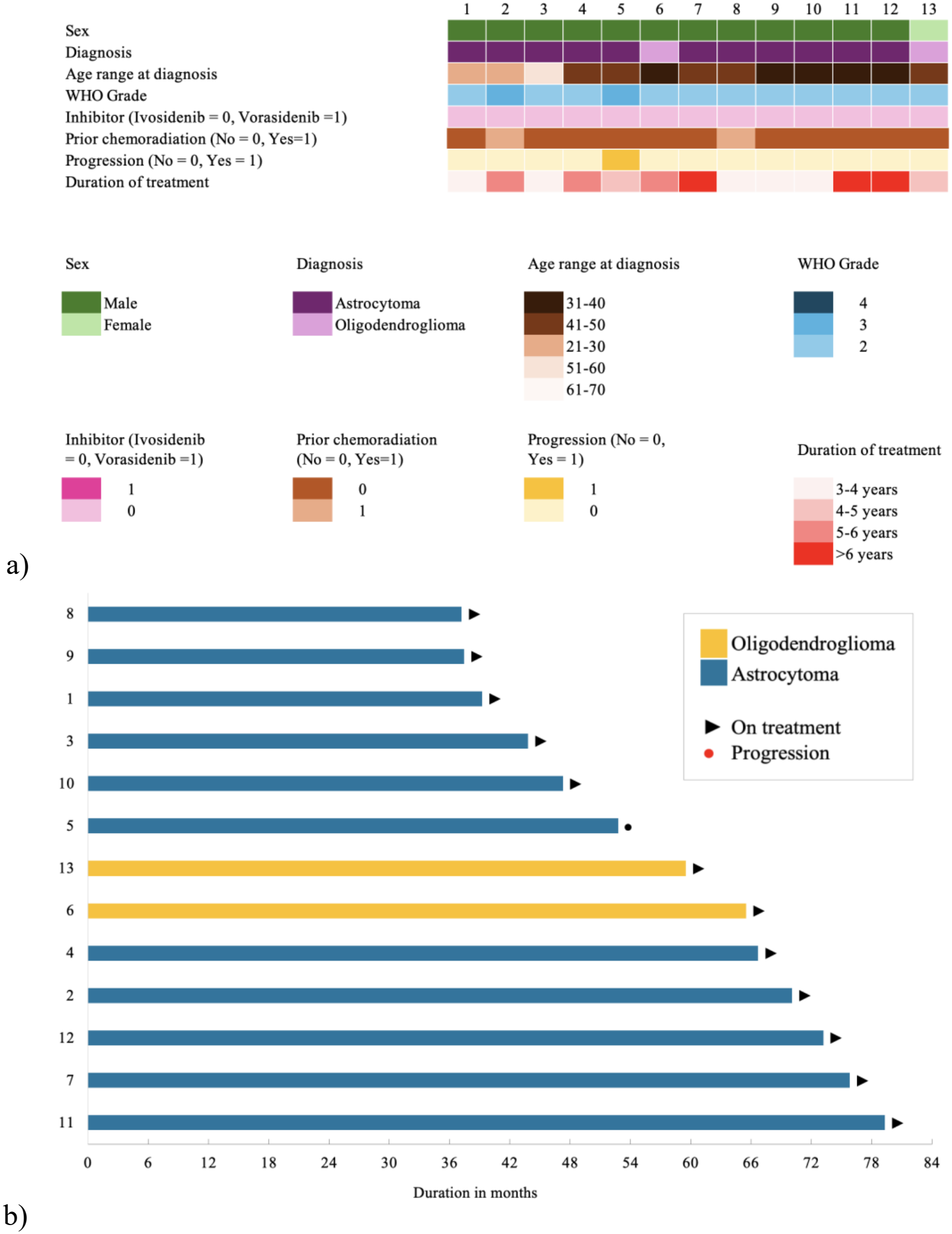
Patient demographics, treatment characteristics, and duration of therapy. **(a)** Oncoplot summarizing patient characteristics, including sex, diagnosis, age at diagnosis, WHO grade of the tumor, prior chemotherapy, use of an IDH inhibitor (ivosidenib), disease progression, and duration of treatment. **(b)** Swimmer plot showing the duration of treatment with ivosidenib for each patient. Arrowheads indicate patients who remained on treatment at the time of data cutoff, whereas red dots indicate disease progression. Bar color denotes tumor histology (oligodendroglioma, yellow or astrocytoma, blue). continued to be on ivosidenib during additional treatments. This patient still met the criteria for stable disease on 3-dimensional measurements at the time of additional therapy **(Supplementary Table 1)**. MRI for patient # 9 beyond the 36-month timeframe was censored since it was performed at an outside facility but was reported to be stable. Other demographic characteristics **(Table 1, Supplementary Figure 1, Supplementary Table 2)** and tumor characteristics **(Supplementary Table 3)** are further summarized.

With a total of 157 MRIs, the mean number of scans evaluated per patient was 9.4 (range 7-15). All patients in this cohort had an IDH1 mutation and were initially started on off-label ivosidenib however 1 patient (13) was changed to vorasidenib mid-treatment, 56 months into therapy because of FDA approval. 2/13 (15%) had a prior history of radiation therapy with chemotherapy (1 with temozolomide, and 1 with PCV) (patient # 2 and patient #8, respectively). 1 patient (patient # 7) was suspected of having radiographic progression 69 months after starting ivosidenib assessed on 2D surveillance scans. Based on tumor board recommendations, this patient was undergoing radiation and temozolomide chemotherapy at the time of this analysis but

**Table 1.** Clinical characteristics of the overall IDH mutant glioma cohort treated with targeted therapy and the subset of long-term responders selected for this study, including sex, age, diagnosis, prior treatments, type of mIDH inhibitor, duration of treatment, and ongoing response status. RT, radiotherapy; PCV, procarbazine, lomustine, and vincristine.

| **Clinical Characteristics** | **Category** | **All patients** | **Final Cohort** |
|---|---|---|---|
| **Patients** |  | 49 | 13 |
| **Sex** | Female | 24/49 | 1/13 |
|  | Male | 25/49 | 12/13 |
| **Age, median (range)** |  | 38 (22-62) | 38 (26-56) |
| **Histopathologic Diagnosis** | Astrocytoma | 32/49 | 11/13 |
|  | Oligodendroglioma | 17/49 | 2/13 |
| **Prior Treatments** | Surgery only | 41 | 11 |
|  | RT+PCV | 2 | 1 |
|  | RT+Temozolomide | 6 | 1 |
| **mIDH Inhibitor Therapy** | Ivosidenib | 46 | 12 |
|  | Vorasidenib | 3 | 0 |
|  | Ivosidenib to Vorasidenib | 1 | 1 |
| **Ongoing responses** |  | 34/49 | 13/13 |
| **Duration of Therapy (years)** |  | 1.96 | 4.96 |

### Longitudinal tumor volume dynamics

Piecewise linear mixed-effects modeling demonstrated distinct temporal phases of radiographic response following initiation of mIDHi therapy.

#### Pretreatment TGR

Prior to treatment, the tumor volume increased significantly at an estimated rate of 12.6 cc/year (β = 12.62, 95% CI 5.51–19.72; P = 0.0007).

#### TGR on treatment

During the first year of therapy, tumor volume declined significantly, with an estimated slope of −9.73 cc/year (β = −9.73, 95% CI −13.81 to −5.64; P < 0.001). After the first treatment year, the slope was no longer significantly different from zero (β = 0.47 cc/year, 95% CI −0.53 to 1.46; P = 0.36), indicating stabilization of tumor volume during prolonged therapy. 9/13 (69%) had an overall reduction in tumor volumes; all had radiographically stable disease with less than 35% volume reduction. However, 4/13 (31%) patients had fluctuating tumor volumes on volumetric measurements with an overall increase but durable control. 3 patients had <20% increase in tumor volumes and 1 had a 43% increase in tumor volumes **(Fig. 3 and Fig. 4, Supplementary Table 4)**.

**Fig 3:**
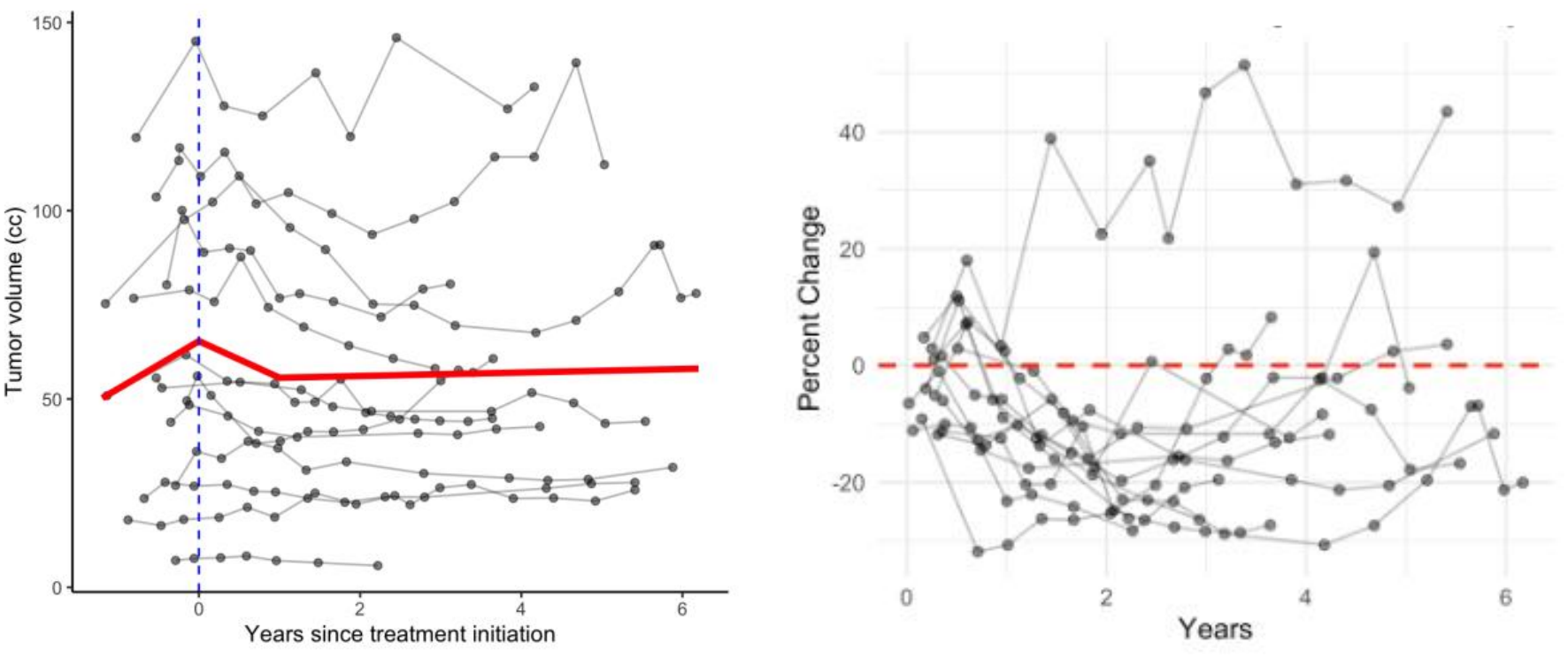
Longitudinal tumor volume dynamics for patients with long term control. Individual longitudinal tumor volume trajectories are shown in absolute volume (cc, left) before and after starting ivosidenib. The percentage changes compared to baseline (right) after starting treatments. Each gray line represents an individual patient with measurements obtained during serial time points of treatment. The red line indicates median tumor growth rate. Treatment initiation is indicated by the dashed vertical line. Tumor volume demonstrated a rapid reduction during the first year of treatment followed by relative stabilization during extended treatment.

**Fig 4:**
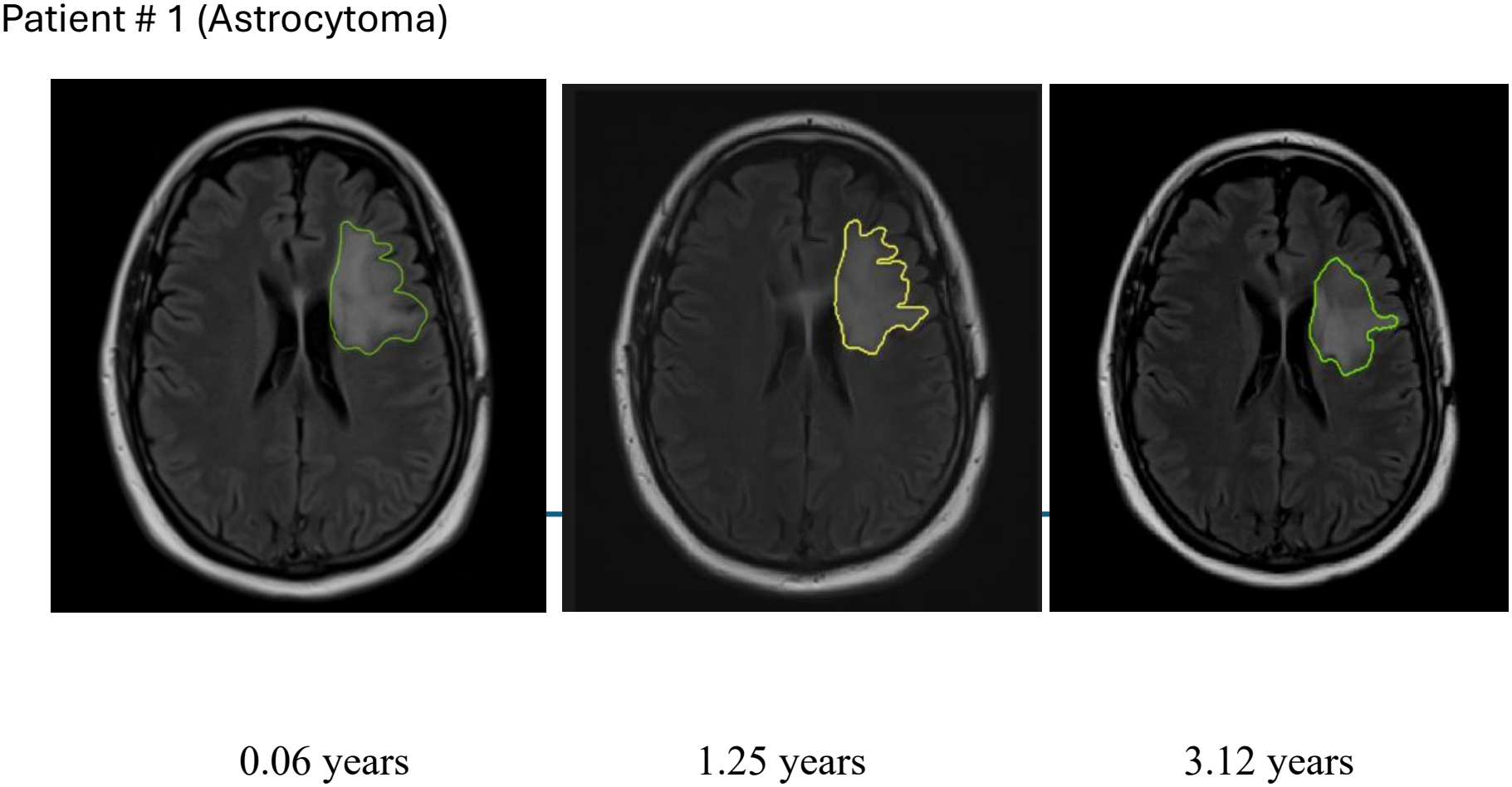
Representative longitudinal T2/FLAIR MRI scans of a patient with Astrocytoma

Overall, the longitudinal mixed-effects analysis demonstrated a biphasic radiographic response characterized by significant tumor shrinkage during the first year of therapy, followed by sustained radiographic stability over long-term treatment without evidence of continued tumor volume reduction or growth.

## Discussion

To our knowledge this is the first, real-world study evaluating the long-term clinical and radiographic outcomes in recurrent IDH mutated glioma treated with targeted therapy. In our study, we observed a reduction in tumor volume within the first year of treatment with ivosidenib followed by stabilization up to and beyond 3 years of treatment. In the entire cohort, approximately two-thirds were on active treatment, consistent with response outcomes observed with clinical trials and prior institutional studies [13, 16, 31]. Interestingly, almost all patients who reached this threshold remained on treatment without later failure, with a median duration of follow-up being 5 years. We selected the 3-year cutoff because at the time of conducting this study, imaging outcomes had been previously reported only up to this time point [18].

Furthermore, all but 1 patient who were able to cross the 36-month timepoint continued to be on treatments suggesting long-term, durable responses. While we only looked at patients who had already crossed the 3-year threshold, 21 patients were still on active treatments who were less than 3 years, and hence the actual percentage of patients with a more than 3-year response might be even higher. Ongoing analysis from INDIGO may confirm our findings and address longer treatment outcomes [18]. Notably, two patients with WHO grade 3 astrocytoma also experienced durable responses despite current guidelines recommending upfront radiotherapy followed by temozolomide based on CATNON [32]. Our results suggest that in recurrent mutant IDH gliomas, targeted inhibition produces early tumor shrinkage followed by prolonged plateau, translating into durable disease control. Thus, the principal benefit of therapy is prolonged disease stabilization with delayed need for chemoradiation in a younger population, hence preserving neurocognitive function, and quality of life [33, 34].

A notable finding in our cohort was the marked heterogeneity in response to treatments, with a subset of patients experiencing prolonged disease control extending beyond 3 years, whereas others progressed substantially earlier. The biological basis for this variability is currently unknown. Although routine genomic profiling is able to identify canonical IDH mutations, it may not fully capture the molecular determinants of long-term sensitivity or acquired resistance to mIDHi therapy. Given that IDH mutation drives widespread epigenetic and metabolic remodeling, differences non-genetic factors may contribute to the observed heterogeneity in treatment response. Future studies integrating longitudinal epigenetic, genomic, metabolic markers, liquid biopsy methods and advanced imaging modalities such as MR Spectroscopy may help identify biomarkers predictive of durable benefit and mechanisms of resistance [35-37]. Lastly, while emerging preclinical data suggests improved radiosensitivity with mIDHi, whether prolonged mutant IDH inhibition ultimately alters the timing of malignant transformation or overall survival largely remains unknown [38, 39].

These findings are significant because the management of IDH-mutant gliomas may increasingly resemble long-term disease control. However, our study has several limitations. Although most patients received ivosidenib rather than vorasidenib, direct comparative efficacy data are lacking. Mellinghoff et al in a phase 1 study found a higher blood brain barrier permeability with vorasidenib however the 2HG reduction were found to be similar with a higher dose of ivosidenib and vorasidenib [12]. Whether the greater CNS penetration of vorasidenib translates into superior long-term clinical outcomes still remains unknown. Likewise, our study was not designed to compare targeted therapy with standard chemoradiation which has been associated with long term sustained reduction in tumor volumes and prospective comparative studies are needed to define the optimal sequencing of these treatment approaches [40]. Second, the retrospective design precluded systematic assessment of patient-reported outcomes, including seizure control, neurocognitive function, and health-related quality of life, which represent important measures of clinical benefit in patients receiving long-term targeted therapy. Third, two patients had previously received radiation and chemotherapy. Although all had progressive disease prior to starting targeted therapy, previous treatment may have altered tumor biology and influenced treatment response. However, inclusion of these patients reflects contemporary clinical practice, in which many individuals initiate targeted therapy after receiving prior treatments. Fourth, despite using three-dimensional volumetric measurements, longitudinal imaging remains susceptible to measurement variability arising from differences in image acquisition, segmentation, and scan quality. This phenomenon, sometimes referred to as the “yo-yo effect,” has been described in prior volumetric studies and may result in fluctuations between response categories despite stable underlying disease [24, 31]. Because the conventional MR imaging-based tumor volumetrics showed limited radiographic changes after the first year of therapy, future studies should evaluate complementary metabolic PET imaging and CSF-based liquid biopsy biomarkers to determine whether they more accurately reflect ongoing treatment response, and emergence of resistance [41-45]. Finally, our analysis was intentionally designed to characterize long-term imaging trajectories among durable responders rather than estimate treatment efficacy or summarize single-center treatment outcomes whereas we addressed early response in our prior report [31]. Consequently, these findings should not be interpreted as the probability of achieving long-term benefit with mIDH inhibition. Despite these limitations, our study provides the longest real-world longitudinal evaluation of mIDHi use, offering remarkable clinical insights for treating oncologists into treatment responses over extended follow-up.

Taken together, our findings suggest that long-term targeted therapies can meaningfully alter the clinical course of progressive IDH-mutant glioma by providing durable disease control, as measured by stable T2/FLAIR volumes, thus allowing a carefully selected patient population to defer initial or salvage chemoradiation for years.

## Data Availability

All data produced in the present work are contained in the manuscript.

## Acknowledgments

Appreciate editorial feedback from John Laterra and Sandra Puetate.

---

**Supplementary Table 1:** Description of Volumetric Response to Treatments.

| **Volumetric Response** |  |
|---|---|
| Complete Response | 100% decrease |
| Partial Response | <100% and > 65% decrease |
| Minor Response | < 65% decrease and >40% decrease |
| Stable Disease | <40% decrease and <40% increase |
| Progressive Disease | >40% increase |

**Supplementary Table 2:** Treatment History.

| Patient # | Age at Diagnosis (y) | Gender (M or F) | Tumor Type (O or A) | Location | Prior Radiation | Prior Chemotherapy (Temozolomide or PCV) | Number of MRI Scans |
|---|---|---|---|---|---|---|---|
| 1 | 26-30 | M | A | Left Frontal | No | No | 9 |
| 2 | 26-30 | M | A | Right Frontal | Y | Y - Tmz | 13 |
| 3 | 56-60 | M | A | Left Frontotemporal | No | No | 11 |
| 4 | 46-50 | M | A | L temporal | No | No | 9 |
| 5 | 41-45 | M | A | Right Frontal | No | No | 7 |
| 6 | 36-40 | M | O | Right Frontoparietal | No | No | 10 |
| 7 | 41-45 | M | A | Left insula | No | No | 14 |
| 8 | 41-45 | M | A | Left Temporal | Y | Y - PCV | 7 |
| 9 | 36-40 | M | A | Left Temporal | No | No | 5 |
| 10 | 31-35 | M | A | Bifrontal | No | No | 11 |
| 11 | 36-40 | M | A | Left frontotemporal | No | No | 7 |
| 12 | 36-40 | M | A | Left Parietal | No | No | 10 |
| 13 | 41-45 | F | O | Left Frontal | No | No | 9 |

**Supplementary Table 3:** Tumor Characteristics.

| Patient # | Pathology | IDH Mutational Status | Grade | P53 | CDKN2A/B | ATRX | 1p/19q |
|---|---|---|---|---|---|---|---|
| 1 | A | IDH1 132S | 2 | Intact | Intact | Loss | Non-Co-deleted |
| 2 | A | IDH1 | 3 | Unavailable | Unavailable | Unavailable | Non-Co-deleted |
| 3 | A | IDH1 R132H | 2 | Mutated | Intact | Lost | Non-Co-deleted |
| 4 | A | IDH1 R132H | 2 | Mutated | Intact | Lost | Non-Co-deleted |
| 5 | A | IDH1 R132H | 3 | Mutated | Intact | Lost | Non-Co-deleted |
| 6 | O | IDH1 | 2 | Unavailable | Unavailable | Intact | Co-deleted |
| 7 | A | IDH1 | 2 | Mutated | Intact | Lost | Non-Co-deleted |
| 8 | A | IDH1 R132H | 2 | Intact | Intact | Lost | Non-Co-deleted |
| 9 | A | IDH1 R132H | 2 | Mutated | Intact | Lost | Non-Co-deleted |
| 10 | A | IDH1 R132C | 2 | Intact | Intact | Lost | Non-Co-deleted |
| 11 | A | IDH1 132G | 2 | Mutated | Intact | Unavailable | Non-Co-deleted |
| 12 | A | IDH1 132G | 2 | Intact | Intact | Loss | Non-Co-deleted |
| 13 | O | IDH1 | 2 | Intact | Intact | Intact | Co-deleted |

**Supplementary Table 4a:** Pre-Treatment Tumor Volume Change Spreadsheet.

| Patient # | Pretreatment MRI |  |  |  |  |  |
|---|---|---|---|---|---|---|
|  | Timepoint -3 (yr) | Tumor Volumes (cc) | Timepoint -2 (yr) | Tumor Volumes (cc) | Timepoint -1(yr) | Tumor Volumes (cc) |
| 1 |  |  | -0.4 | 80.31 | -0.21 | 100.08 |
| 2 | -0.88 | 17.88 | -0.47 | 16.38 | -0.19 | 18.01 |
| 3 |  |  | -1.14 | 50.9 | -0.16 | 61.7 |
| 4 |  |  | -0.53 | 55.6 | -0.46 | 52.94 |
| 5 |  |  | -0.35 | 43.83 | -0.12 | 48.41 |
| 6 | -0.68 | 23.59 | -0.42 | 27.9 | -0.06 | 26.86 |
| 7 |  |  | -1.16 | 75.3 | -0.18 | 97.62 |
| 8 |  |  | -0.81 | 76.74 | -0.12 | 78.95 |
| 9 |  |  | -0.29 | 7.13 | -0.06 | 7.67 |
| 10 |  |  | -0.15 | 49.53 | -0.02 | 56.08 |
| 11 |  |  | -0.78 | 119.41 | -0.04 | 144.99 |
| 12 |  |  | -0.29 | 27.03 | -0.03 | 36.07 |
| 13 | -0.53 | 103.64 | -0.25 | 113.28 | -0.24 | 116.7 |

**Supplementary Table 4b:**
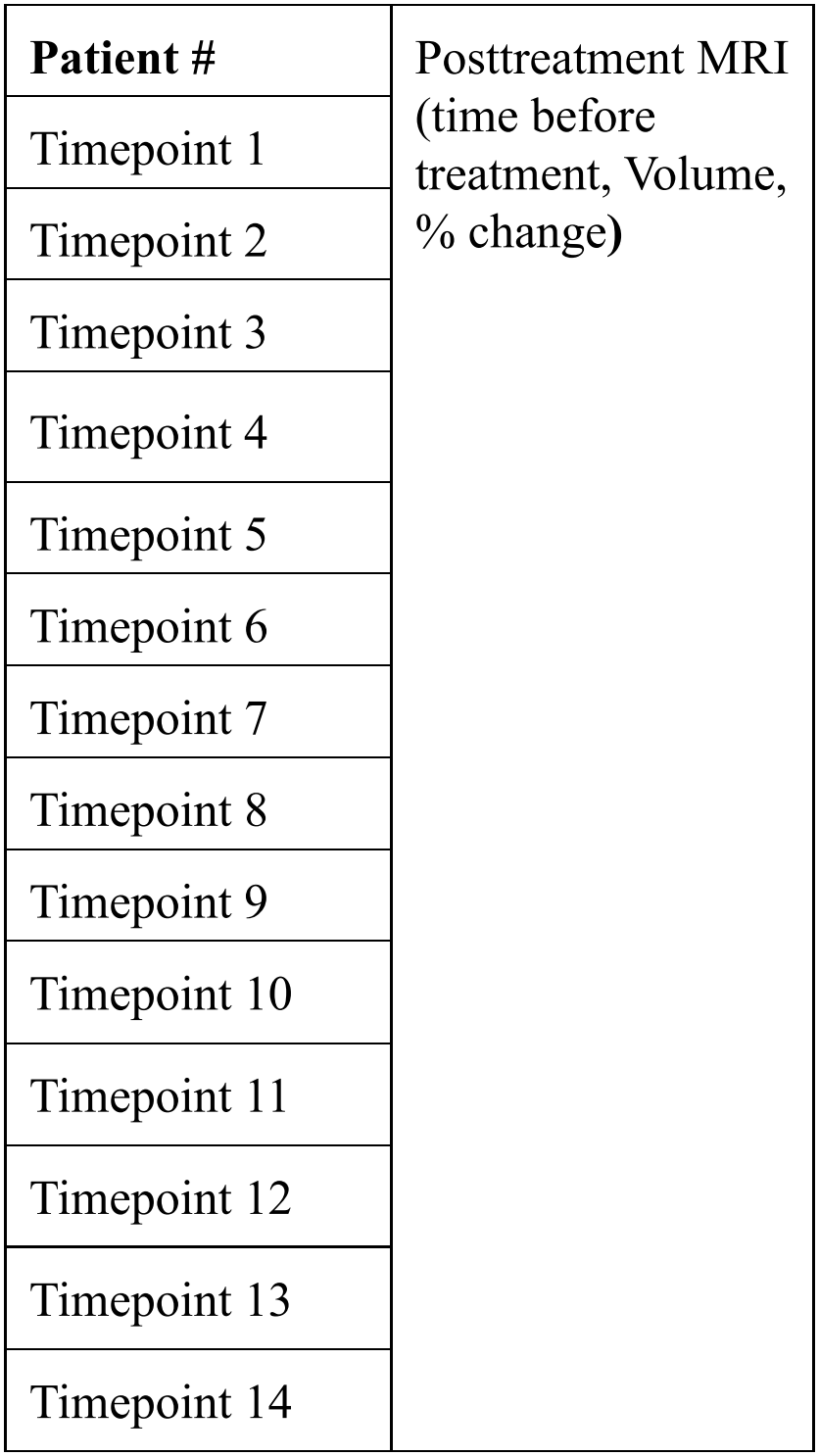
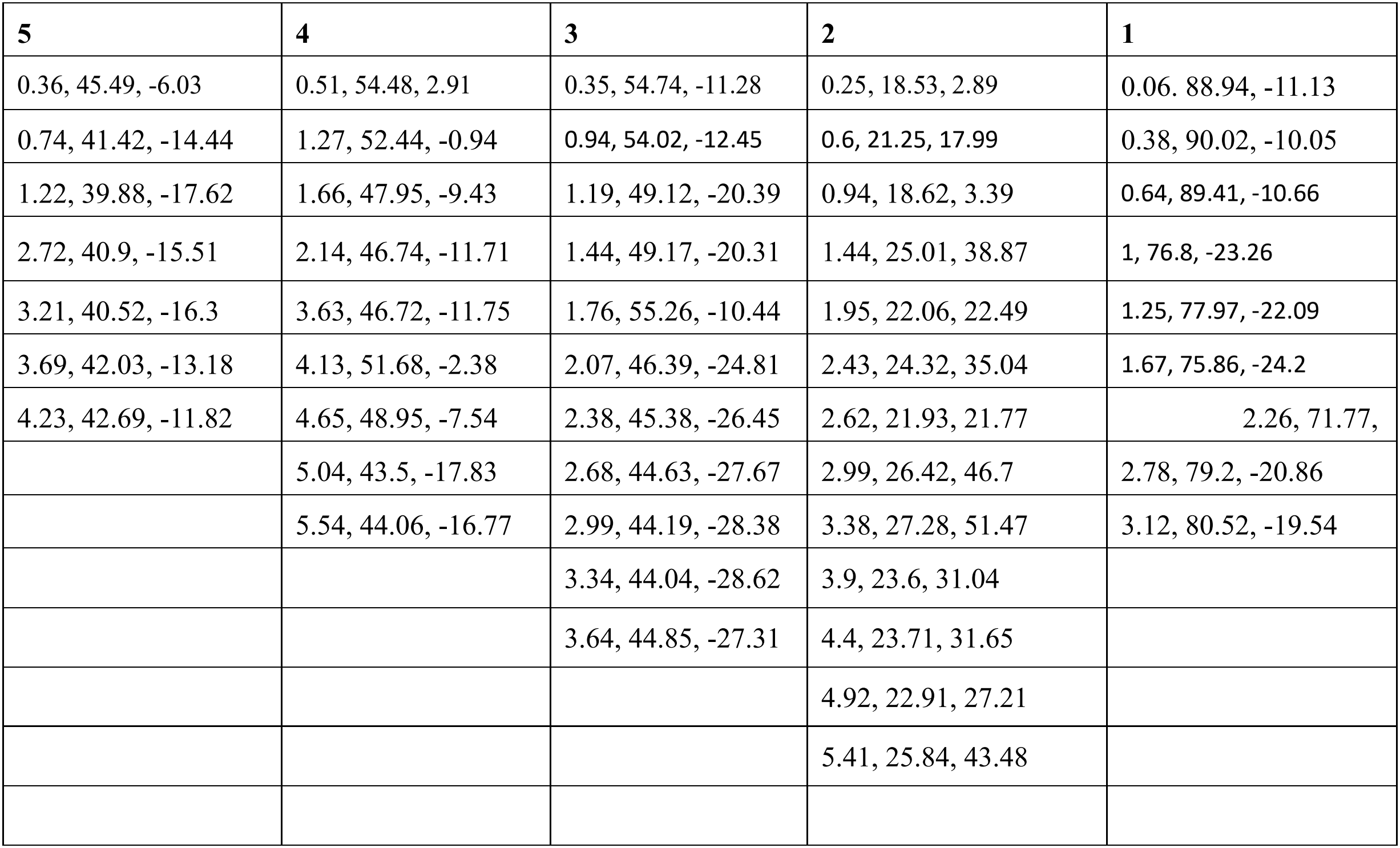
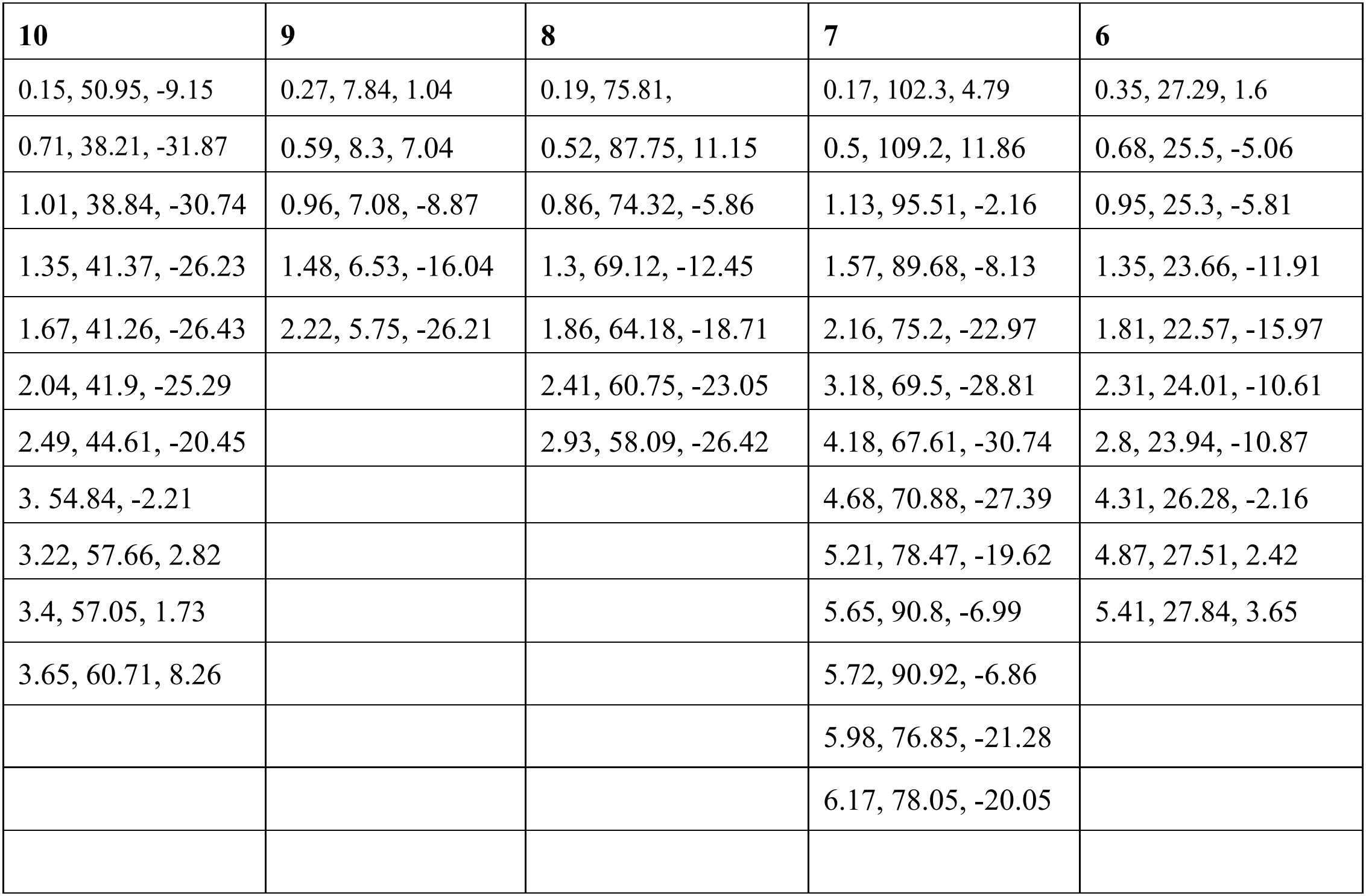
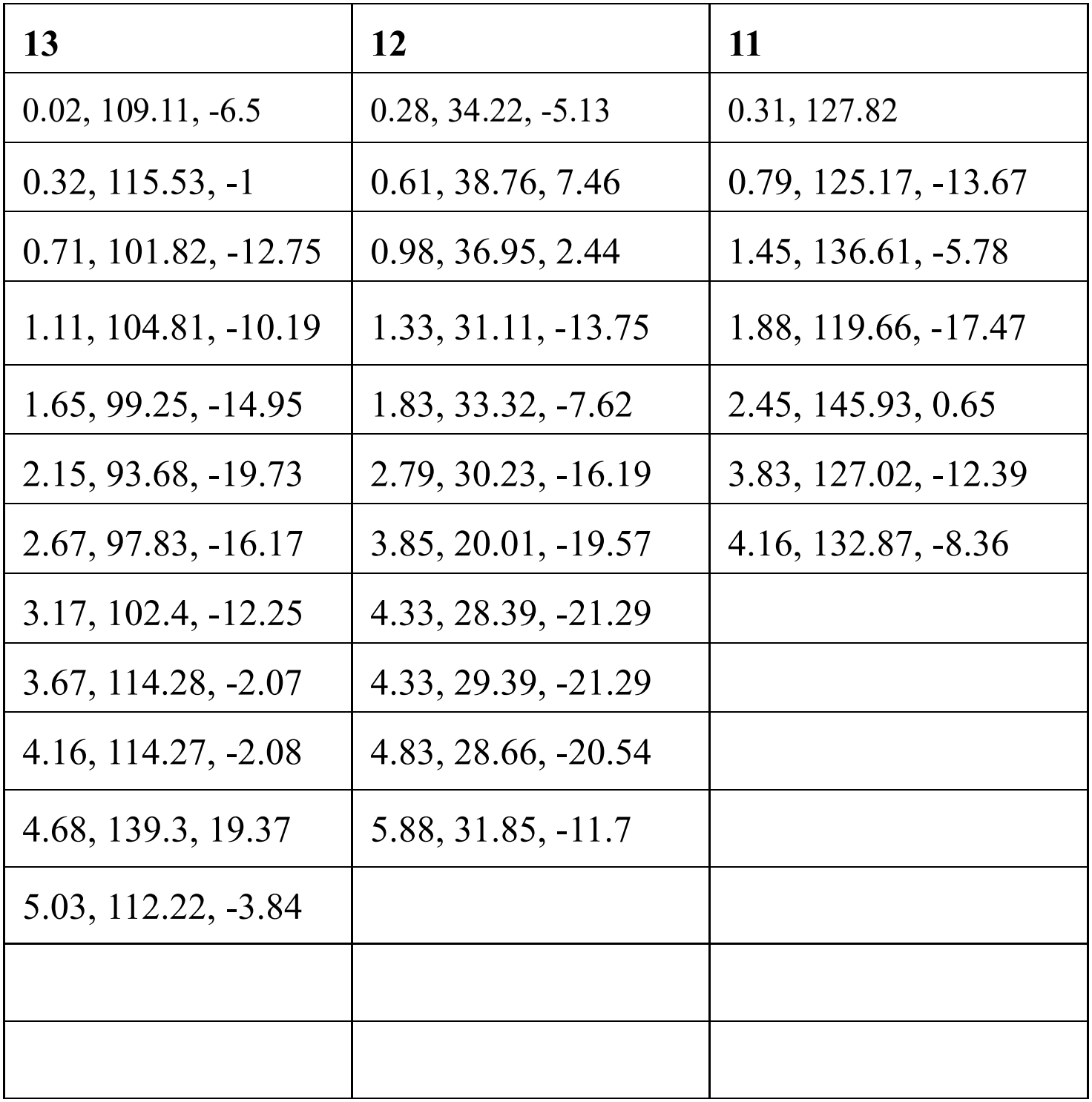
Post-Treatment Tumor Volume Change Spreadsheet.

| **Patient #** | Posttreatment MRI (time before treatment, Volume, % change**)** | **5** | **4** | **3** | **2** | **1** | **10** | **9** | **8** | **7** | **6** | **13** | **12** | **11** |
|---|---|---|---|---|---|---|---|---|---|---|---|---|---|---|
| Timepoint 1 |  | 0.36, 45.49, -6.03 | 0.51, 54.48, 2.91 | 0.35, 54.74, -11.28 | 0.25, 18.53, 2.89 | 0.06. 88.94, -11.13 | 0.15, 50.95, -9.15 | 0.27, 7.84, 1.04 | 0.19, 75.81, | 0.17, 102.3, 4.79 | 0.35, 27.29, 1.6 | 0.02, 109.11, -6.5 | 0.28, 34.22, -5.13 | 0.31, 127.82 |
| Timepoint 2 |  | 0.74, 41.42, -14.44 | 1.27, 52.44, -0.94 | 0.94, 54.02, -12.45 | 0.6, 21.25, 17.99 | 0.38, 90.02, -10.05 | 0.71, 38.21, -31.87 | 0.59, 8.3, 7.04 | 0.52, 87.75, 11.15 | 0.5, 109.2, 11.86 | 0.68, 25.5, -5.06 | 0.32, 115.53, -1 | 0.61, 38.76, 7.46 | 0.79, 125.17, -13.67 |
| Timepoint 3 |  | 1.22, 39.88, -17.62 | 1.66, 47.95, -9.43 | 1.19, 49.12, -20.39 | 0.94, 18.62, 3.39 | 0.64, 89.41, -10.66 | 1.01, 38.84, -30.74 | 0.96, 7.08, -8.87 | 0.86, 74.32, -5.86 | 1.13, 95.51, -2.16 | 0.95, 25.3, -5.81 | 0.71, 101.82, -12.75 | 0.98, 36.95, 2.44 | 1.45, 136.61, -5.78 |
| Timepoint 4 |  | 2.72, 40.9, -15.51 | 2.14, 46.74, -11.71 | 1.44, 49.17, -20.31 | 1.44, 25.01, 38.87 | 1, 76.8, -23.26 | 1.35, 41.37, -26.23 | 1.48, 6.53, -16.04 | 1.3, 69.12, -12.45 | 1.57, 89.68, -8.13 | 1.35, 23.66, -11.91 | 1.11, 104.81, -10.19 | 1.33, 31.11, -13.75 | 1.88, 119.66, -17.47 |
| Timepoint 5 |  | 3.21, 40.52, -16.3 | 3.63, 46.72, -11.75 | 1.76, 55.26, -10.44 | 1.95, 22.06, 22.49 | 1.25, 77.97, -22.09 | 1.67, 41.26, -26.43 | 2.22, 5.75, -26.21 | 1.86, 64.18, -18.71 | 2.16, 75.2, -22.97 | 1.81, 22.57, -15.97 | 1.65, 99.25, -14.95 | 1.83, 33.32, -7.62 | 2.45, 145.93, 0.65 |
| Timepoint 6 |  | 3.69, 42.03, -13.18 | 4.13, 51.68, -2.38 | 2.07, 46.39, -24.81 | 2.43, 24.32, 35.04 | 1.67, 75.86, -24.2 | 2.04, 41.9, -25.29 |  | 2.41, 60.75, -23.05 | 3.18, 69.5, -28.81 | 2.31, 24.01, -10.61 | 2.15, 93.68, -19.73 | 2.79, 30.23, -16.19 | 3.83, 127.02, -12.39 |
| Timepoint 7 |  | 4.23, 42.69, -11.82 | 4.65, 48.95, -7.54 | 2.38, 45.38, -26.45 | 2.62, 21.93, 21.77 | 2.26, 71.77, | 2.49, 44.61, -20.45 |  | 2.93, 58.09, -26.42 | 4.18, 67.61, -30.74 | 2.8, 23.94, -10.87 | 2.67, 97.83, -16.17 | 3.85, 20.01, -19.57 | 4.16, 132.87, -8.36 |
| Timepoint 8 |  |  | 5.04, 43.5, -17.83 | 2.68, 44.63, -27.67 | 2.99, 26.42, 46.7 | 2.78, 79.2, -20.86 | 3. 54.84, -2.21 |  |  | 4.68, 70.88, -27.39 | 4.31, 26.28, -2.16 | 3.17, 102.4, -12.25 | 4.33, 28.39, -21.29 |  |
| Timepoint 9 |  |  | 5.54, 44.06, -16.77 | 2.99, 44.19, -28.38 | 3.38, 27.28, 51.47 | 3.12, 80.52, -19.54 | 3.22, 57.66, 2.82 |  |  | 5.21, 78.47, -19.62 | 4.87, 27.51, 2.42 | 3.67, 114.28, -2.07 | 4.33, 29.39, -21.29 |  |
| Timepoint 10 |  |  |  | 3.34, 44.04, -28.62 | 3.9, 23.6, 31.04 |  | 3.4, 57.05, 1.73 |  |  | 5.65, 90.8, -6.99 | 5.41, 27.84, 3.65 | 4.16, 114.27, -2.08 | 4.83, 28.66, -20.54 |  |
| Timepoint 11 |  |  |  | 3.64, 44.85, -27.31 | 4.4, 23.71, 31.65 |  | 3.65, 60.71, 8.26 |  |  | 5.72, 90.92, -6.86 |  | 4.68, 139.3, 19.37 | 5.88, 31.85, -11.7 |  |
| Timepoint 12 |  |  |  |  | 4.92, 22.91, 27.21 |  |  |  |  | 5.98, 76.85, -21.28 |  | 5.03, 112.22, -3.84 |  |  |
| Timepoint 13 |  |  |  |  | 5.41, 25.84, 43.48 |  |  |  |  | 6.17, 78.05, -20.05 |  |  |  |  |
| Timepoint 14 |  |  |  |  |  |  |  |  |  |  |  |  |  |  |

